# Emerging Mental Health Challenges, Strategies and Opportunities in the context of the COVID-19 Pandemic: Perspectives from South American Decision-makers

**DOI:** 10.1101/2020.07.16.20155630

**Authors:** Daniel A. Antiporta, Andrea Bruni

## Abstract

**Background:** Mental health awareness has increased during the COVID-19 pandemic. Although international guidelines address the mental health and psychosocial support (MHPSS) response to emergencies, regional recommendations on COVID-19 are still insufficient. We identified emerging mental health problems, strategies to address them, and opportunities to reform mental health systems during the COVID-19 pandemic in South America.

**Methods:** An anonymous online questionnaire was sent to mental health decision-makers of Ministries of Health in 10 South American countries in mid-April 2020. The semi-structured questionnaire had 12 questions clustered into 3 main sections: emerging challenges in mental health, current and potential strategies to face the pandemic, and, key elements for mental health reform. We identified keywords and themes for each section through summative content analysis.

**Findings:** An increasing mental health burden and emerging needs are arising as direct and indirect consequences of the pandemic among health care providers and the general population. National lockdowns challenge the delivery and access to mental health treatment and care. Strategies to meet these health needs rely heavily on timely and adequate responses by strengthened mental health governance and systems, availability of services, virtual platforms, and appropriate capacity building for service providers. Short- and medium-term strategies focused on bolstering community-based mental health networks and telemedicine for high-risk populations. Opportunities for long-term mental health reform entail strengthening legal frameworks, redistribution of financial resources and collaboration with local and international partners.

**Interpretation:** Mental health and psychosocial support have been identified as a priority area by South American countries in the COVID-19 response. The pandemic has generated specific needs that require appropriate actions including: implementing virtual based interventions, orienting capacity building towards protection of users and health providers, strengthening evidence-driven decision making and integrating MHPSS in high-level mechanisms guiding the response to COVID-19.

**Funding:** None.

**Research in context:** *Evidence before this study:* The COVID-19 pandemic has affected mental health and wellbeing as well as its determinants. General population have reported anxiety and stress while health professionals fear, and bereavement. Mental health services have also been overburdened as the health needs increase as consequence of the pandemic and the isolation measures in place. The WHO General director has recognized mental health and psychological support (MHPPS) as a major pillar in the overall health response to the COVID-19 pandemic. Likewise, the Inter Agency Standing Committee (IASC) published a global briefing recommending eight MHPPS interventions to be implement during the crisis. Nonetheless, evidence to guide action at regional and sub-regional levels is still insufficient.

*Added value of this study:* This study provides expert perspectives of decision-makers about mental health burden and actions during the COVID-19 in South America, currently the most serious hub of infection worldwide. Health services have reported an increase of anxiety, stress and fear among the general population emerging during the pandemic. The pandemic has generated specific needs that require appropriate actions including implementing virtual based interventions, bolstering community-based mental health networks, and integrating MHPSS in high-level mechanisms guiding the response to COVID-19. Decision-makers identified opportunities to seize for long-term mental health reform such as strengthening legal frameworks, redistribution of financial resources and collaboration with local and international partners.

*Implications of all the available evidence:* The importance of this research goes beyond documenting the status quo of mental health at country level, but implies fostering, enhancing and expanding collaborations in the Sub-region to strengthen the mental health response to the COVID-19 pandemic. Country-cooperation initiatives in mental health have been an important strategy to improve local mental health systems and services. Our findings are expected to better orient next steps in making decisions on mental health policies and services in South America, but also to inform public health key leaders and mental health experts within and beyond the Region of the Americas.

## I. Introduction

Mental health and psychosocial problems are expected to rise during adversity and crisis(1), such as the COVID-19 pandemic(2), but awareness of mental health has already increased in media and academic platforms(3). General population have reported anxiety and stress(4, 5) while health professionals and frontline aid workers reported fear and bereavement(6). Isolation measures, discontinuity in health services, and scarce availability of medications represent additional barriers to preserving a good mental health.

Mental health and psychosocial support (MHPSS) have been recognized as major components within the overall health response to the COVID-19 pandemic(7). MHPSS include strategies to protect or promote psychological well-being and prevent mental conditions. The Inter Agency Standing Committee (IASC) has provided guidelines to address mental health and psychosocial aspects during the epidemic(8). While the IASC provides guidance on a global level, evidence to guide action at regional and sub-regional levels is still insufficient. Region and country-based research can help reduce the evidence-gap on local mental health action and strategies.

Historically mental health care has been severely under-resourced; however, some regions like South America have made substantial progress regarding national policies and legal frameworks. For instance, Peru initiated a radical mental health reform in 2015 that produced a shift from a hospital-centered mental health towards a community-based model(9). In 2019 Paraguay was identified as the only country in the Region of the Americas to participate in the WHO Mental Health Special Initiative, which aims to ensure universal health coverage for mental health(10).

The SARS-CoV-2 virus has spread widely in South America with 1,119,575 confirmed cases and 48,860 deaths as of June 7^th^, 2020(11). Recent forecasting models suggest a dramatic scenario for the coming months projecting substantial increase in cases and deaths by August(12), despite wide lockdowns and curfews implemented in several countries. While many South American countries are fiercely fighting against the spread of the virus, additional challenges, for example increased mental health needs, have arisen amidst this pandemic. Key responses in mental health entail a deeper understanding of local needs and interventions as well as identification of opportunities to strengthen mental health care.

The relevance of this study lies in the need to generate evidence on the radical changes and emerging challenges created by the COVID-19 pandemic. We gathered information from decision makers on the main needs in terms of mental health and wellbeing in South America. We identified emerging mental health challenges and strategies to reduce the negative impact of the epidemics, and, key opportunities to reform mental health systems and services by seizing opportunities during the crisis.

## II. Methods

### Sample

We identified mental health focal points in Ministries of Health in all 10 countries that belong to the Pan American Health Organization (PAHO) South American subregion: Argentina, Bolivia, Brazil, Chile, Colombia, Chile, Paraguay, Peru, Uruguay, and Venezuela. Electronic invitations were sent by email to at least one focal point per country between the April 10-15, 2020. Eligible participants were 1) at least 18 years old, and, 2) holding a high-level managerial position within their mental health directions or units.

### Procedures

An anonymous online questionnaire was designed in Qualtrics (Provo, UT) about mental health in the context of the COVID-19 pandemic. The questionnaire (see Table 1) had 9 country-specific questions, divided in three sections: 1) emerging challenges in mental health, 2) current and potential strategies (short-term, next two months, and medium-term, next six months, to face the pandemic, and, 3) strategies and opportunities for mental health reform.

**Table I.**
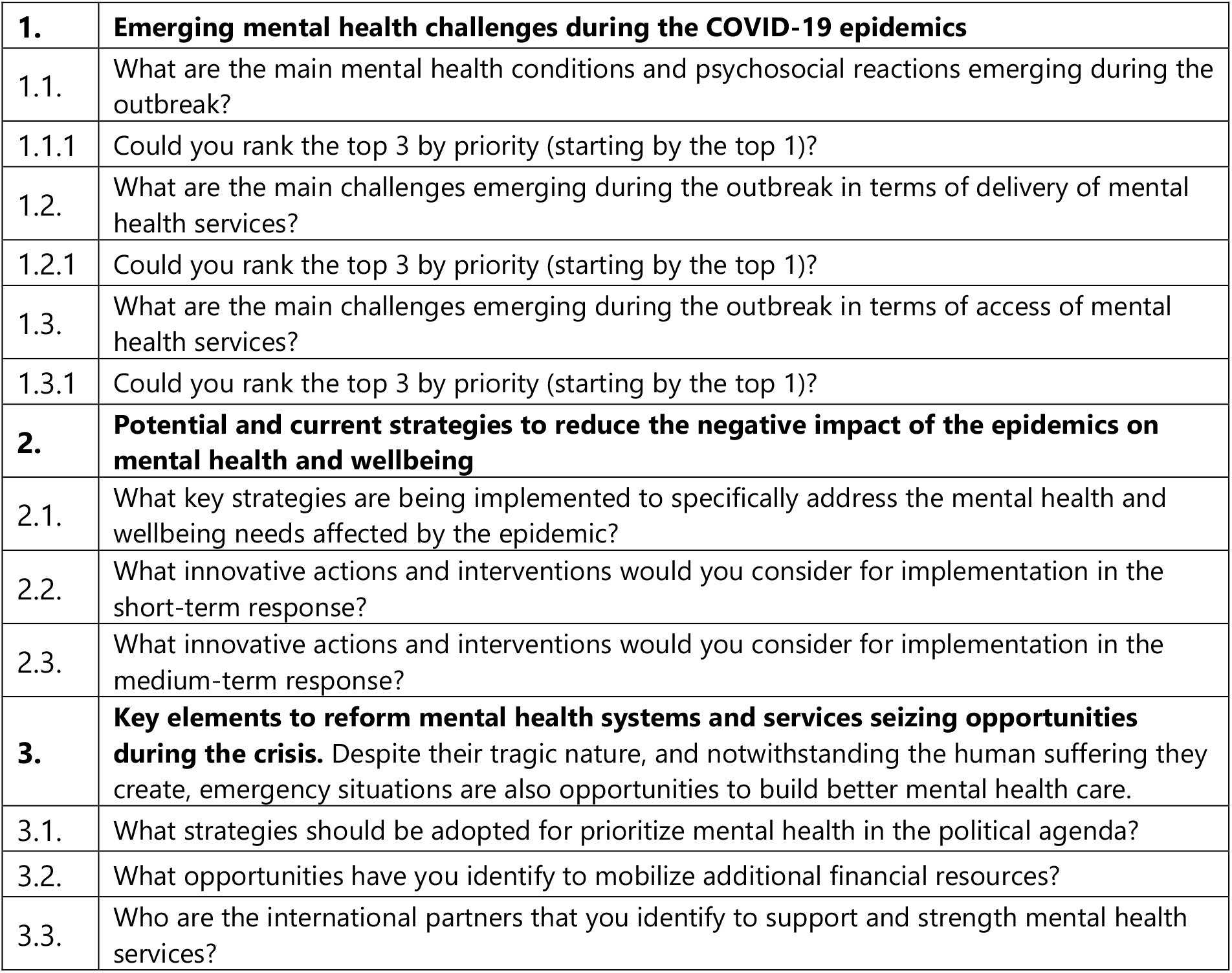
Semi-structured Questionnaire for Directors/staff of Mental Health Units.

|  |  |
| --- | --- |
| **1.** | **Emerging mental health challenges during the COVID-19 epidemics** |
| 1.1. | What are the main mental health conditions and psychosocial reactions emerging during the outbreak? |
| 1.1.1 | Could you rank the top 3 by priority (starting by the top 1)? |
| 1.2. | What are the main challenges emerging during the outbreak in terms of delivery of mental health services? |
| 1.2.1 | Could you rank the top 3 by priority (starting by the top 1)? |
| 1.3. | What are the main challenges emerging during the outbreak in terms of access of mental health services? |
| 1.3.1 | Could you rank the top 3 by priority (starting by the top 1)? |
| **2.** | **Potential and current strategies to reduce the negative impact of the epidemics on mental health and wellbeing** |
| 2.1. | What key strategies are being implemented to specifically address the mental health and wellbeing needs affected by the epidemic? |
| 2.2. | What innovative actions and interventions would you consider for implementation in the short-term response? |
| 2.3. | What innovative actions and interventions would you consider for implementation in the medium-term response? |
| **3.** | **Key elements to reform mental health systems and services seizing opportunities during the crisis.** Despite their tragic nature, and notwithstanding the human suffering they create, emergency situations are also opportunities to build better mental health care. |
| 3.1. | What strategies should be adopted for prioritize mental health in the political agenda? |
| 3.2. | What opportunities have you identify to mobilize additional financial resources? |
| 3.3. | Who are the international partners that you identify to support and strength mental health services? |

The questionnaire had a semi-structured format and was designed to last around 20 minutes to complete. Following ethical principles, all participants voluntarily consented to participate in this questionnaire. The protocol and questionnaire were submitted to the Johns Hopkins Bloomberg School of Public Health and the PAHO ethics review committee and all procedures were exempted for review.

### Data Analysis

A qualitative approach was used performing summative content and thematic analysis. The research team first read the raw data of 2 questionnaires and generated an initial codebook. The codebook and themes were revised and updated based on gaps shown by the initial list. Early versions of findings were jointly reviewed by the team to agree on interpretability of results. Analytical products are themes and keywords by each section. We used the software ATLAS.ti 8 for Windows to facilitate the data management and organization.

### Working definitions

For this study, we have used the following terms considering the local context and the purposes of the study. We have used the terms *lockdown, quarantine*, and *home-stay policies* interchangeably. While most participants referred to home-stay policies or lockdowns, they used the term “*quarantine*” in all cases to describe these policies. By stress reactions, we intended to capture all mental and psychosocial conditions and reactions that require ‘any type of support that aims to protect or promote psychosocial wellbeing and/or prevent or treat mental disorder(13). This included keywords such as stress, post-traumatic stress, acute stress, and severe stress. Lastly, we used anxiety to describe terms such as anxiety disorders and anguish.

### Role of the funding source

There was no funding source for this study. The corresponding author and first author had full access to all the data and had final responsibility for the decision to submit for publication.

## III. Results

We received back 9 out of 10 complete questionnaires representing 9 countries, and 44% of respondents were female. The median time for completion was 55 minutes (IQR=170 min). All respondents worked in high ranked decision-making positions in mental health units, programs, or departments in their ministry of health of their respective countries.

### Emerging mental health problems and psychosocial reactions

Informants reported up to 12 different emerging problems across countries. The most frequent mental health and psychosocial reactions reported were anxiety (12 mentions), stress (8 mentions), and fear (4 mentions). Reactions were attributed not only to the pandemic itself but also to public health measures that countries implemented to control the disease, such as total lockdowns and home-stay policies. Another common topic was domestic violence affecting children and women. Less frequent problems included insomnia, irritability, solitude, and sadness, especially among those who are living alone.

Informants were asked to rank the top 3 different emerging problems based on the urgency to intervene. Anxiety or anxiety disorders were ranked top across informants, although few also indicated stress reactions and fear. The second highest problem was not equally homogenous across informants, and answers ranged from an increase in consumption of substances, to stress reactions and depression. The third top priority was heterogenous and included domestic violence, substance use and impulsive reactions.

### Challenges for mental health services delivery

Participants indicated several challenges for mental health care delivery arising during the COVID-19 pandemic, for which the identification of high-risk populations is necessary to plan appropriate responses. Prioritized target populations include service providers, patients who were already in contact with the mental health services and potential new users that might need mental health support as a result of the pandemic, including those people that lost a relative or a loved one. The focus on health providers should not be limited to emergency room (ER) and intensive care unit (ICU) health professionals but should include mental health professionals as well.

Challenges that referred to the services offered were grouped into outpatient services, inpatient care, and availability of medications. Outpatient services challenges included the limited capacity of health services to use virtual/telemedicine platforms to provide care to specific populations, i.e., elderly people, and indigenous communities, or to disseminate key messages and relevant information through mass media. Challenges for inpatient care concerned adequate time for admissions and care provided during the lockdown. Disruption in availability of psychotropic medications was described in terms of reduced access and distribution to inpatient and outpatient care facilities.

Other challenges related to organizational interventions and training for health providers. Participants referred to the need for an action plan to strengthen community based mental health services and bridge the mental health treatment gap. They also referred to the need for synergies with public institutions and civil society to strengthen public mental health surveillance, interventions, and communication. Additional challenges were the limited availability of virtual platforms and limited time for training service providers on adequate responses of mental health care, such as psychological first aid.

The challenge that ranked highest was to maintain or reopen primary mental health care services to adequately respond to the needs of affected people and overcome the limitations of providing MHPSS interventions during the lockdown. Other topics included the training of mental health providers, distribution of medications and caring for mental health providers. The second highest challenge was heterogeneous across participants: some referred to caring for the mental health and wellbeing of frontline workers, and adequate functioning of inpatient care. Challenges ranked as the highest third referred to establishing or strengthening intersectoral work and providing psychosocial support to people and families affected by COVID-19.

### Challenges for mental health services access

The main reported barriers for accessing mental health services, including therapies and other types of care, are the national lockdown measures, which have shut down most primary health centers to stop spreading the disease. Scarce resources to reorganize mental health services to virtual forms and systems for appointments were also described as challenges that jeopardize access to services. The delivery of virtual-based treatments and interventions relies on the availability of services as well as patient’s expertise to use technological tools, which are not optimal during the current scenario.

Access to care is also reduced due to the limited number of professional and functioning community centers with mental health care available during the pandemic. Access to medications was also reported as a potential challenge given the lower availability of psychotropic medications as compared to those used in general health and for conditions related to the COVID-19.

The highest ranked challenge was the continuity of care using strategies that respect the lockdown measures and providing appropriate care for patients. Another topic that ranked highly was the limited availability of trained mental health providers in the primary care level. As second top challenges, participants described limited access to psychotropic medications during the pandemic as well as the lack of training and resources to implement telemedicine sessions. The third top challenge mentioned by participants was reaching out to vulnerable populations, such as those with low income. Another topic was the activation of emergency services for mental health to respond to increased demand during this crisis.

### Strategies to face the needs in mental health during the pandemic

Strategies that have been implemented frequently during this time included the use of mass communication media, at national and local levels by Ministries of Health and community health centers. Communications were tailored by life course stage or ethnicity in some countries. Ongoing efforts aim to promote self-help mechanisms through social networks, at the national, regional, and municipal level.

Other strategies included establishing or strengthening mental health services through telemedicine and national or local hotlines for mental health care and psychosocial support. Some countries reported hotlines for specific populations such as the elderly or people with disabilities. Additional use of virtual platforms was referred by some respondents to implement mental health training, the exchange of experiences between territories and reporting to stakeholders from the national and local levels.

A common short-term strategy was to ensure the adequate mental care of admitted persons, their families, and health providers at psychiatric hospitals as well of those in the highest risk units (i.e. ER and ICU). Special attention was given to those patients who might need to be admitted in a context of limited availability of beds. Psychosocial support was considered a crucial strategy to prevent stress reactions due to burnout and other consequences of the pandemic among health providers.

Access to pharmacological treatment was also a concern during the early stages. Some strategies proposed to bridge the gap included the establishment of a virtual delivery system and partnerships with existing pharmaceutical networks to facilitate the access to these treatments. Communication strategies for psychosocial education and support considering cultural and gender perspectives were also suggested. Examples of the latter is the digital system of mental health care for health providers in Chile.

For the medium-term, strong relevance was given to the creation and use of virtual platforms and applications for delivery of mental health services, strengthening of social networks and offering psychosocial support. A mobile application in Colombia that aims to screen for COVID-19 symptoms, was cited as an example of an app that can provide mental health care information to a wider population. The need for a community-based mental health system, which strengthens the capacities of non-specialized primary health providers, was a recurrent topic among respondents.

Participants also mentioned the elaboration or strengthening of protocols and programs to provide care to people positive for COVID-19, specific populations, through the adequate implementation of the Mental Health Gap Action Programme (mhGAP) in the areas most affected by the crisis. Participants also referred to the need for mental health professionals to be included in the multidisciplinary team that provides care to people affected by COVID-19.

### Opportunities for Mental Health Reform

The increased awareness of mental health by stakeholders, media, and the general population, during the pandemic and the lockdowns, represents an opportunity to increase visibility of mental health, to mobilize resources and to prioritize mental health policies and interventions. Communication strategies through social media and official channels can highlight the importance of mental health by offering self-help messages to manage stress and other reactions during the lockdown period. Training health care providers in mental health strategies can increase awareness among these professionals.

Effective advocacy and leadership need to focus on strengthening mental health planning and legislation to adequately respond to the pandemic. The creation and inclusion of commissions for mental health within technical working groups will allow mental health services to be prioritized not only in the current response but also in the post-pandemic scenario. Partnerships with local organizations and civil society are key to enhance the role of mental health response during the crisis.

Reorienting mental health services towards a community-based system will provide appropriate care tailored to the needs of the population. Moreover, building information systems using timely and robust data will allow the monitoring of mental health burden and associated factors, to better inform stakeholders.

When asked about opportunities to mobilize additional financial resources for mental health, respondents had few suggestions based on their country experience. Some suggested redirecting resources from more specialized facilities towards the primary health networks or community-based centers. Effective collaborations with local and regional authorities might facilitate the implementation of current mental health policies and lead to allocation of additional funds.

International partners, such as cooperation bodies, were considered as potential sources of financial support and collaboration for articulated efforts in mental health care during the pandemic. The most cited institutions were PAHO/WHO (8/9 respondents) and UNICEF (3/9 respondents). On average, each participant reported at least 3 institutions.

## IV. Discussion

An emerging burden of mental health needs is arising as direct and indirect consequences of the pandemic among the general population as well as health care providers in South America. National lockdowns and social distancing measures challenge the delivery and access of mental health care and treatment. Strategies to meet these health needs heavily rely on timely and adequate responses by strengthened mental health governance and systems, availability of services and virtual platforms and appropriate capacity building for service providers. Short- and medium-term strategies focus on the implementation of community-based mental health systems, virtual support, communications, and appropriate care for populations in vulnerable or high-risk settings. Opportunities for long-term mental health reform entail strengthening legal frameworks, redistribution of financial resources and collaboration with local and international partners.

Situations of emergencies normally translates into higher prevalence of mental conditions, including stress reactions, common and severe mental disorders(1, 14). Participants reported a higher burden of anxiety and stress in their respective countries. Other reactions can be also expected in severe COVID-19 infections such as fatigue, delirium, and neuropsychiatric syndromes(15). Detrimental effects in mental health have been reported in the general population(4, 5), health providers(16-18) as well as overburdening health services(19).

The novel cohabitation circumstance, consequence of lockdowns and home-stay policies, may represent an opportunity for exchange among family members and loved ones, but at the same time may result in increased tensions(20, 21) and violence(2), including violence against women and children. Home confinement has increased the prevalence of depressive symptoms among children(22). People with mental health disorders and disabilities may suffer further disruption in services and accommodation prior to COVID-19(23). Increased consumption of alcohol and psychotropic substances will turn a difficult situation into a more challenging scenario. Mental health plays a pivotal role in this context since it has plenty to contribute to improving positive coping mechanisms to face new challenges and hardships Participants emphasized the relevance of ensuring the continuity of services. The capacity to adjust to the increased volume of people in need largely depends on the previous mental health infrastructure. Importantly, health systems with well-developed community-based mental health networks are more likely to adjust to the novel scenario. Conversely, health systems that are centered in acute care hospitals and psychiatric hospitals will struggle to respond to the increasing needs of the population and the pre-existing mental health treatment gap may become more dramatic. The Ministry of health of Peru, for instance, is taking measures to ensure the community-based mental health centers keep functioning in a virtual manner, when possible, and by face-to-face interventions, if needed. The community-based mental health centers represent an invaluable resource at the heart of mental health community networks to respond to the needs of the population.

Another important feature that emerged was put in evidence during the COVID-19 pandemics is the need for expanding mental health services through telemedicine initiatives. The risk of infection of service users and health professionals, among others, can be reduced through the implementation of virtual-based interventions, whenever possible. Successful examples of these interventions have been shown in China, where they implemented surveys, mental health education and psychological counselling services though online services(24). In Colombia, a smartphone application was launched to provide support to COVID-19 affected populations; this application includes a component entirely dedicated to mental health that orient users and provides them with appropriate information(25). Maintaining a limited proportion of face-to-face interventions seems to be crucial, provided that sufficient protective measures are always taken to protect service users and health professionals. Those settings where community-based services are more developed will be better positioned also to reorient the methods of delivering services and ‘go virtual’.

Situations of crisis, such as the COVID-19 pandemic, despite its inherent disruptive dramatic consequences, may generate important opportunities for improving mental health services(26). All respondents in our study presented their insights in seizing potential opportunities to reform mental health services in a long-term perspective. Given the special nature of COVID-19 and its profound impact on mental health and wellbeing of populations, this long-term approach seems fundamental in providing insights to decision makers beyond the most immediate response to the critical event.

This study identified decision makers in high rank positions of mental health units at Ministries of Health as key informants during the early stage of the pandemic in South America. Besides sharing their expert opinion, this group was instrumental in identifying specific strategies to operationalize recommendations. Despite participants being high-ranking officials, the survey was directed to a limited number of individuals. Thus, the authors acknowledge the need for generating additional evidence and further investigate the perspectives of relevant key actors, including: senior mental health professionals, such as psychiatrists and psychiatric nurses, but also representatives from civil society such as service users and their family members or caregivers. We gathered information from all countries except for Brazil, the biggest and most affected country by COVID-19 pandemic in the subregion(11), due to administration changes during the time of data collection. Nevertheless, our aim was to portray a sub-regional situation in mental health challenges and not country-specific profiles, for which collecting information from decision-makers of 9 out of 10 countries allowed us to fulfill the objective.

Mental health and psychosocial support have been identified as a priority area by South American countries in the COVID-19 response. The pandemic has generated specific needs that require appropriate actions including: implementing virtual based interventions, orienting capacity building towards protection of users and health providers, strengthening evidence-driven decision making and integrating MHPSS in high-level mechanisms guiding the response to COVID-19. The results of this study are expected to better orient next steps in making decisions on mental health policies and services in South America, but also to inform public health key leaders and mental health experts within and beyond the Region of the Americas.

## Data Availability

The de-identified data that support the findings of this study are available from the corresponding author, AB, upon reasonable request.

## Acknowledgements

The authors acknowledge the participants of this study, their disposition to contribute to our research and the mental health responses they are leading in their countries.

## Notes

### Competing Interest Statement

The authors have declared no competing interest.

### Funding Statement

No external funding was received for this study.

### Author Declarations

The PAHO ethics review committee (PAHOERC) and the JHSPH Institutional Review Board considered this study as non human subjects research and was exempted.

